# Genetics and Epigenetics of Aldehyde Dehydrogenase (ALDH2) in Alcohol Related Liver Disease

**DOI:** 10.1101/2021.04.16.21255566

**Authors:** Bhagyalakshmi Shankarappa, Jayant Mahadevan, Pratima Murthy, Meera Purushottam, Biju Viswanath, Sanjeev Jain, Harshad Devarbhavi, Ashok V. Mysore

**Affiliations:** St John’s Medical College Hospital, Bangalore, India; National Institute of Mental Health and Neurosciences (NIMHANS), Bangalore, India

## Abstract

Alcohol dependence and cirrhosis are key outcomes of excessive alcohol use. We studied the interaction between genetics and epigenetics at the aldehyde dehydrogenase (*ALDH2*) locus to understand differences in vulnerability to cirrhosis. Individuals were selected according to ICD 10 criteria for Alcohol dependence with Cirrhosis (AUDC+ve, N=116) and Alcohol dependence but without Cirrhosis; (AUDC-ve, N=123) from the clinical services of Gastroenterology and Psychiatry at the St John’s Medical College Hospital (SJMCH). Fibroscan/sonographic evidence was used to rule out fibrosis for the AUDC-ve group. Genomic DNA from blood was used for genotyping at *ALDH2* (rs2238151) locus. A subset of the samples was assessed for DNA methylation (AUDC+ve, N=50; AUDC-ve, N=50) at the *LINE-1* and *ALDH2* CpG loci by pyrosequencing on a PyroMark Q24.

*LINE1* DNA methylation did not differ between the groups. *ALDH2* DNA methylation was significantly lower in AUDC+ve group compared to AUDC-ve group (P <0.001). Lower methylation in T-allele carriers compared to T-allele non-carriers of the *ALDH2* locus (rs2238151) was observed in AUDC+ve subjects (P=0.009). Compromised methylation in blood DNA at candidate loci, in those with liver disease in the context of prolonged severe alcohol abuse, could be explored as a biomarker for current pathology, and further progression.

## Introduction

According to the World Health Organisation, 2.3 billion people consume alcohol in the world, and about 75 million are classified as having alcohol use disorders. The harmful use of alcohol results in 3 million deaths worldwide and 5.1% of disability adjusted life years (DALY) globally (World Health Organization, 2019). A substantial proportion of alcohol attributable burden of disease comes from two conditions namely, alcohol use disorders (AUD) and alcohol related liver disease (ARLD) which make up for one quarter of alcohol attributable mortality and one third of alcohol attributable disease burden measured in DALYs (Rehm and Shield, 2019). Given that the liver is the primary site of ethanol metabolism, it is prone to tissue injury by heavy drinking (Mathurin and Bataller, 2015).

The pathogenesis of ARLD involves alcohol-metabolizing enzymes that alter the levels of acetaldehyde and Reactive Oxygen Species (ROS). The genes that code for these enzymes show considerable allelic variation, worldwide. Liver cirrhosis is a complex disease. The onset, progression, and clinical outcome of liver cirrhosis are influenced by alleles in candidate genes that are often associated with particular ethnic backgrounds, as well as by environmental factors. These genetic and environmental factors may influence the metabolism of alcohol, and nutrients, and contribute to the pathobiology of liver disease. Additionally, genes involved in immune regulation, insulin sensitivity, oxidative stress and extracellular matrix deposition may also modulate the degree of severity (Ramos-Lopez et al., 2015).

ARLD is a spectrum, with alcohol induced steatosis seen in more than 90% of individuals who drink heavily and regularly. Among them, about 10 to 20 % progress to alcoholic steatohepatitis, that is thought to be a precursor to cirrhosis (Bruha et al., 2012). In general, the mechanisms postulated for the development of alcoholic steatohepatitis are oxidative stress (from alcohol metabolism and reactive oxygen species formation), immune dysregulation, and endotoxins from the gut (Ceni et al., 2014). However, the reasons behind individual predilection towards these conditions are not well understood. While factors like age, gender, diet and quantity/frequency of alcohol use are important, a significant role may be played by genetic and epigenetic factors (Meroni et al., 2018).

Ethanol is oxidised to acetaldehyde by alcohol dehydrogenase (ADH), and subsequently to acetate by aldehyde dehydrogenase (ALDH). ALDH is expressed in a wide range of tissues with the highest expression noted in the liver (Human protein atlas). There are two major ALDH isoforms, cytosolic and mitochondrial, encoded by the *ALDH1* and *ALDH2* genes, respectively. Of the two isoforms, mitochondrial ALDH2 plays the central role in human acetaldehyde metabolism because of its submicromolar Km for acetaldehyde (<1 µM) (Jiang et al., 2020). In addition to its role in elimination of acetaldehyde, ALDH also plays a critical role in the detoxification of lipid peroxidation products which are generated due to oxidative stress (Nene et al., 2017).

It is well established that the rs671 variant in *ALDH2*, that is found almost exclusively among East Asians, results in reduced aldehyde dehydrogenase activity and is associated with a lower risk of AUD (Edenberg and McClintick, 2018). It has also been seen that on alcohol exposure in East Asians, the rs671 variant in *ALDH2* is associated with a greater risk of esophageal, hepatocellular carcinomas (Li et al., 2016), adverse cardiovascular outcomes (Cook et al., 2020) and cognitive decline (Chen et al., 2019).

However, there are a number of other variants in *ALDH2*, that are common in other parts of the world and may also affect the activity of the enzyme (Chen et al., 2020). The SNP studied in the present study for *ALDH2* (rs2238151), has been previously reported to be associated with delayed acetaldehyde catabolism and level of alcohol consumption (Dickson et al., 2006; Hakenewerth et al., 2011).

Consumption of excessive amounts of alcohol is also known to influence global DNA methylation, though studies have shown inconsistent findings (Zhang and Gelernter, 2017). Our earlier work looking at global DNA methylation using LINE1 (Long-Interspersed Nuclear Elements-1) loci showed hypomethylation in patients with AUD compared to controls (Soundararajan et al., 2021).

Alcohol has a toxic effect on many tissues, and the different responses may culminate in organ-specific disease syndromes such as cirrhosis, neuropathy, and neuropsychiatric manifestations. Although the metabolism of alcohol is relatively well studied, a greater understanding of genetic factors, and their overlap with DNA methylation, can help us to explain the how and why of alcohol related end organ complications.

In this study we evaluated the DNA methylation pattern of LINE1 and ALDH2 CpG loci in AUD patients with and without cirrhosis. We also investigated the distribution of the *ALDH2* rs2238151 polymorphism in the two groups.

## Materials and Methods

### Samples

Patients from the outpatient or inpatient services, of Gastroenterology and Psychiatry at St John’s Medical College Hospital (SJMCH), who met the International Classification of Diseases (ICD-10) criteria for Alcohol Dependence with Cirrhosis (AUDC+ve; N=116), and gave consent, were invited to participate. Competing causes of liver disease such as hepatitis B, hepatitis C and NAFLD were excluded. Another group of patients who had Alcohol Dependence without cirrhosis (AUDC-ve; N=123) were also studied for comparison. The study was approved by the institutional ethics committee. Those included had no past history of mental illness themselves, or in their first-degree relatives. The AUDC-ve group were those who were drinking heavily for a prolonged period, but with no clinical suspicion of alcohol induced steatohepatitis or cirrhosis. Fibroscan (Liver Stiffness Measurement, LSM 7-12.5 kPa, moderate to severe fibrosis; LSM>12.5 likely-Cirrhosis) and/or sonographic findings (enlarged spleen, ascites, low blood platelets and severe gastrointestinal (GI) bleeding from dilated blood vessels (called varices) that can rupture) were used to rule out fibrosis in AUDC-ve group.

### Genotyping

The DNA was extracted using Qiagen DNA isolation Kit (DNAeasy Blood and Tissue Kit) from the blood samples collected from AUDC+ve and AUDC-ve subjects. Genotyping for *ALDH2* rs2238151 polymorphism was done by PCR-RFLP method (Roy et al., 2016). PCR was performed in a 20 µl reaction volume containing 50mM Tris buffer with MgCl2, 2.5mM each dNTP, DNA Taq polymerase. The PCR product (size 598bp) was incubated with 0.1U HaeIII restriction enzyme for 2 h. The restricted fragments were electrophoresed on 2% agarose gel. Digestion of the PCR product in genotype TT yielded 598 bp, CT yielded 598 bp, 382 bp and 215 bp while CC genotype had 382 bp and 215 bp products.

### DNA Methylation

A randomly selected subset of 100 subjects (AUDC+ve=50; AUDC-ve=50) was processed for estimation of DNA methylation using Pyrosequencing. Firstly, 450 ng of DNA was subjected to bisulfite conversion (Zymo gold Bisulfite Kit). Bisulfite converted DNA was amplified using Pyromark PCR kit (Qiagen, Valencia, CA) with primers specific for LINE-1 repetitive elements (for global methylation) (Choi et al., 2009) and the *ALDH2* CpG loci. A predesigned pyromark assay (Qiagen) was used for ALDH2 DNA methylation analysis including 5 CpG sites. The biotin labeled PCR products were immobilized using streptavidin sepharose beads (GE LifeSciences, Uppsala, Sweden) and sequenced by pyrosequencing on the Q24 System (Qiagen) according to manufacturer’s instructions. Methylation (%5-mC) was calculated using Pyro Q-CpG Software (Qiagen). We examined a single cytosine at a non-CpG site within PCR products to ensure successful bisulfite conversion of unmethylated cytosines. We used the averaged value of the 5 CpG sites of LINE1 to estimate global DNA methylation.

### Statistical analysis

Hardy Weinberg equilibrium was assessed using software (http://www.oege.org/software/hwe-mr-calc.shtml). All statistical analyses were performed using R version 4.0.2. Based on the presence or absence of the risk allele ‘T’, different genetic models were used. Presence of risk allele ‘T’ was coded as ‘1’ and absence as ‘0’. All continuous variables have been presented as mean and standard deviation values. Methylation values have been expressed as a percentage of methylated cytosines over the total of methylated and unmethylated cytosines. Independent t-test was used to compare mean methylation levels and methylation at individual CpG sites. Pearson correlations were run for analysing correlation and partial correlations for controlling variables. The P value <0.05 was considered to be statistically significant.

## Results

### Sociodemographic and Clinical Details

Demographic and clinical characteristics of participants are described in Table 1. The two groups did not differ in age, but those in the AUDC+ve group had used lesser quantities of alcohol (Mean ± SD = 13.1±6.4 units/day) compared to the AUDC-ve group (Mean ± SD = 15.3 ±7.1units/day) (P = 0.01). They had also been drinking for a comparatively shorter duration (16.01±7.2 years vs 17.33±7.1 years), and had a later age of onset of drinking (29.5±8.8 years vs 24.5±7.4 years) (p < 0.001). As expected, the AUDC+ve had higher levels of conjugated bilirubin, direct bilirubin, AST/ALT ratio and ALP and GGT, and lower levels of total protein and albumin, as well as lower haemoglobin levels and Mean Corpuscular Volume when compared to those in the AUDC-ve group (Table 1).

**Table 1.** Comparison of sociodemographic, clinical variables and biochemical parameters between individuals with AUD with and without cirrhosis in the entire sample and DNA methylation subset

| Study Variable (Mean±SD) | Whole sample set (N=239) |  |  | Subset for DNA Methylation Study (N=100) |  |  |
| --- | --- | --- | --- | --- | --- | --- |
|  | AUDC-ve(N=123) | AUDC+ve(N=116) | P-value | AUDC-ve(N=50) | AUDC+ve(N=50) | P-value |
| **Age** | 42.08±10.3 | 45.21±9.0 | **0.01** | 44.25±11.3 | 43.25±8.7 | 0.6 |
| **Years of Drinking** | 17.33±7.14 | 16.01±7.2 | 0.13 | 19.78±8.2 | 12.09±5.14 | **<0.001** |
| **Alcohol intake in Units** | 15.3±7.1 | 13.1±6.4 | **0.01** | 14.8±7.14 | 13.6±6.4 | 0.4 |
| **Age at onset of Drinking** | 24.5±7.4 | 29.5±8.8 | **<0.001** | 24.06±7.8 | 30.7±8.7 | **<0.001** |
| **Total Bilirubin** | 1.32 ± 1.06 | 8.70 ± 9.00 | **<0.001** | 1.32 ± 1.27 | 9.43 ± 9.90 | **<0.001** |
| **Direct Bilirubin** | 0.58 ± 0.64 | 5.71 ± 6.55 | **<0.001** | 0.60 ± 0.87 | 6.17 ± 6.62 | **<0.001** |
| **AST** | 131.98 ± 155.91 | 108.25 ± 104.54 | 0.15 | 103.62 ± 90.19 | 126.11 ± 135.75 | 0.33 |
| **ALT** | 77.61 ± 62.37 | 49.24 ± 46.22 | **<0.001** | 70.49 ± 41.80 | 58.08 ± 57.06 | 0.2 |
| **AST/ALT Ratio** | 1.71 ± 1.12 | 2.36 ± 1.22 | **<0.001** | 1.47 ± 0.97 | 2.35 ± 1.27 | **<0.001** |
| **GGT** | 382.17 ± 417.52 | 266.88 ± 380.04 | **0.02** | 361.13 ± 460.40 | 246.20 ± 298.47 | 0.14 |
| **ALP** | 114.79 ± 60.35 | 162.74 ± 111.69 | **<0.001** | 108.73 ± 45.94 | 182.63 ± 154.65 | **0.001** |
| **GGT/ALP Ratio** | 3.38 ± 3.23 | 1.64 ± 2.10 | **<0.001** | 3.11 ± 3.05 | 1.43 ± 1.67 | **<0.001** |
| **Total Protein** | 7.18 ± 0.71 | 6.56 ± 1.01 | **<0.001** | 7.40 ± 0.55 | 6.45 ± 0.97 | **<0.001** |
| **Albumin** | 3.81 ± 0.76 | 2.46 ± 0.73 | **<0.001** | 3.67 ± 0.64 | 2.59 ± 1.79 | **<0.001** |
| **Hemoglobin** | 14.55 ± 2.34 | 9.35 ± 2.53 | **<0.001** | 14.18 ± 2.2 | 10.16 ± 2.8 | **<0.001** |
| **MCV** | 95.92 ± 7.33 | 92.86 ± 10.06 | **0.005** | 95.39 ± 7.87 | 95.97 ± 8.65 | 0.72 |

Legend: *Bil* – Bilirubin – Normal range 0.1 to 1.2 mg/dL; *AST* - Aspartate Aminotransferase – Normal range 5-40 U/L; *ALT* - Alanine aminotransferase - Normal range 7-56 U/L; *AST/ALT ratio* - Normal range <1; *GGT* – Gamma- Glutamyl Transferase (Normal range 9-48 U/L); *ALP* – Alkaline Phosphatase - Normal range 20-140 IU/L; *GGT/ALP* - Normal range <1; *WBC*- White Blood Cells (Normal value 4.5 to 11.0 × 10<sup>9</sup>/L)

### Differences in genotype and allele frequency between groups

A total of 239 individuals (AUDC+ve, N=116; AUDC-ve, N=123) were genotyped for *ALDH2* rs2238151. The genotype and allele frequency distribution for SNP rs2238151 in AUDC+ve and AUDC-ve groups are shown in Table 2. We observed a trend for the ‘T’ allele frequency to be higher in the AUDC+ve group than the AUDC-ve group, but this was not statistically significant (Table 2). The genotype and allele frequency distribution for SNP rs2238151 in AUDC+ve and AUDC-ve groups showed no significant differences in the sample subset used for DNA methylation (Supplementary Table 1). Clinical variables such as quantity of drinking, years of drinking and age of onset of drinking did not differ significantly based on the presence or absence of ‘T’ allele.

**Table 2.** Differences between Genotype and Allele frequency of ALDH2 rs2238151 in individuals with AUD with and without cirrhosis in the entire sample

| Gene and SNP | Genotype Frequency | AUDC-ve<br>N=123<br>(n, %) | AUDC+ve<br>N=116<br>(n, %) | P | Chi-Square |
| --- | --- | --- | --- | --- | --- |
| ALDH2<br>rs2238151 | CC | 70(0.56) | 53(0.45) | 0.1 | 3.8 |
|  | CT | 47(0.38) | 52(0.44) |  |  |
|  | TT | 6(0.04) | 11(0.09) |  |  |
|  | Allele frequency | 2n=246 | 2n=232 | P | Chi-Square |
|  | C | 0.76 | 0.68 | 0.06 | 3.3 |
|  | T | 0.23 | 0.32 |  |  |

### Comparison of DNA methylation at LINE 1 and ALDH2 loci between groups

LINE 1 DNA methylation levels were not significantly different between AUDC+ve and AUDC-ve groups (Figure1, Supplementary Table 2).

**Figure 1:**
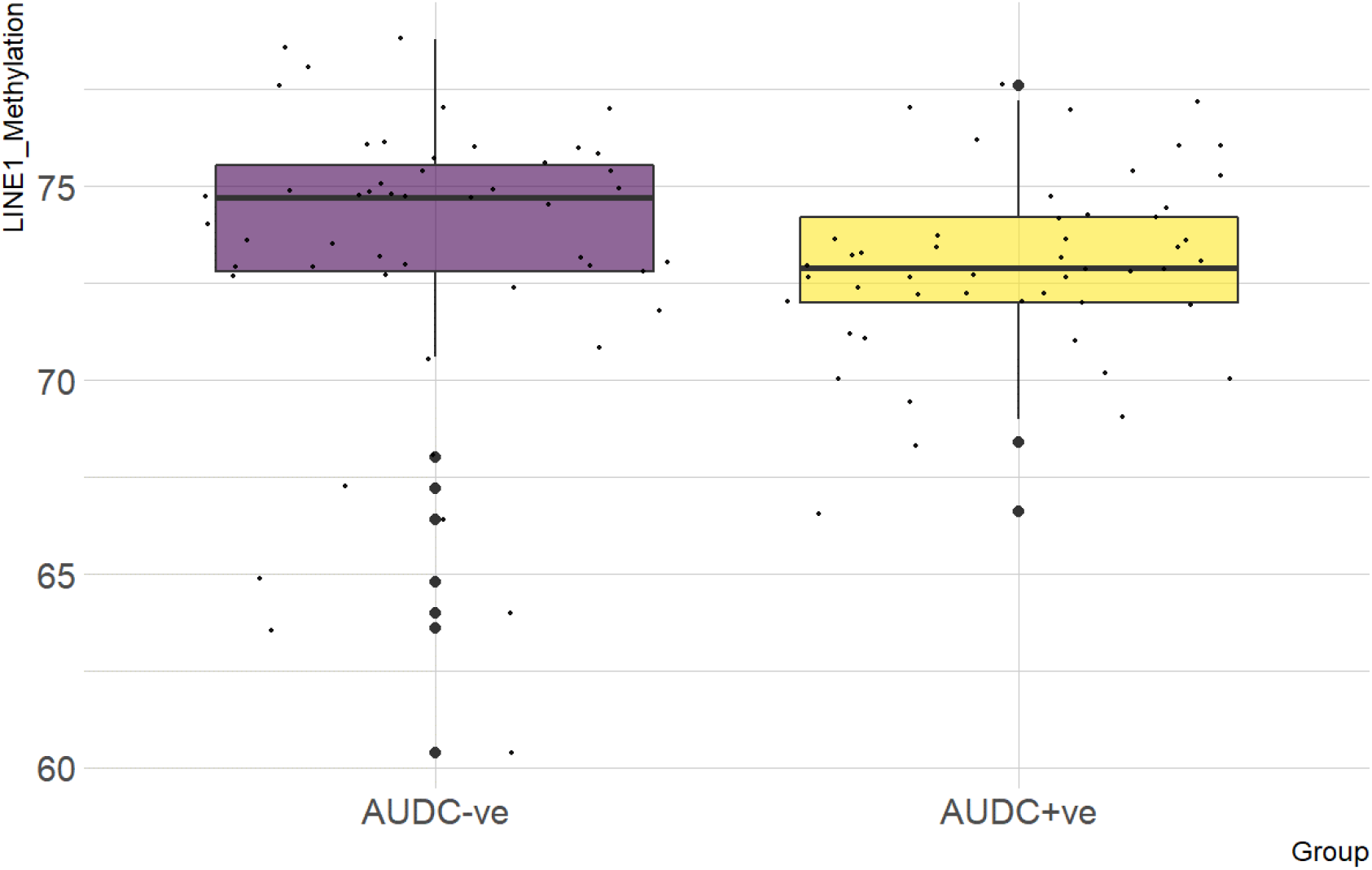
Comparison of mean LINE 1 methylation levels in individuals with AUD with and without cirrhosis. The results indicate that there are no significant differences in LINE 1 methylation in individuals with AUD with and without cirrhosis. Legend: AUDC-ve – Individuals with AUD without cirrhosis, AUDC+ve – Individuals with AUD with cirrhosis

For the *ALDH2* loci, AUDC+ve group had lower levels of mean methylation (3.8±0.8) compared to the AUDC-ve group (5.1±1.9; P<0.001) (Figure 2, Table 3). When we analysed effect of rs2238151 allele in AUDC+ve group, we observed significantly lower methylation in T-allele carriers (3.5±0.7) compared to T-allele Non-carriers (4.0±0.8) (P=0.01) (Table 4). This genotype influence was not seen in the AUDC-ve group or when the entire sample was taken together (Table 4).

**Table 3:** Differences in average DNA methylation of ALDH2 gene CpG sites in individuals with AUD with and without cirrhosis

| ALDH2 gene CpG sites | Methylation Levels (Mean±SD) |  | p-value |
| --- | --- | --- | --- |
|  | AUDC-ve<br>N=50 | AUDC+ve<br>N=50 |  |
| CpG1 | 5.7±2.2 | 3.9±1.2 | <0.001 |
| CpG2 | 3.4±1.9 | 1.8±0.8 | <0.001 |
| CpG3 | 6.1±2.3 | 4.2±1.1 | <0.001 |
| CpG4 | 3.3±2.2 | 1.7±0.7 | <0.001 |
| CpG5 | 8.2±2.6 | 6.8±1.2 | <0.001 |
| Average | 5.1±1.9 | 3.8±0.8 | <0.001 |

**Table 4:** Influence of ALDH2 rs2238151 genotypes on average ALDH2 gene DNA methylation in individuals with AUD with and without cirrhosis

| Groups | Methylation Levels (Mean±SD)<br>based on ALDH2 rs2238151 genotype |  | p-value |
| --- | --- | --- | --- |
|  | CC | CT+TT |  |
| AUDC-ve | 5.0±1.9 | 5.1±1.9 | 0.86 |
| AUDC+ve | 4.0±0.8 | 3.5±0.7 | 0.01 |
| Total | 4.6±1.6 | 4.2±1.6 | 0.23 |

**Figure 2:**
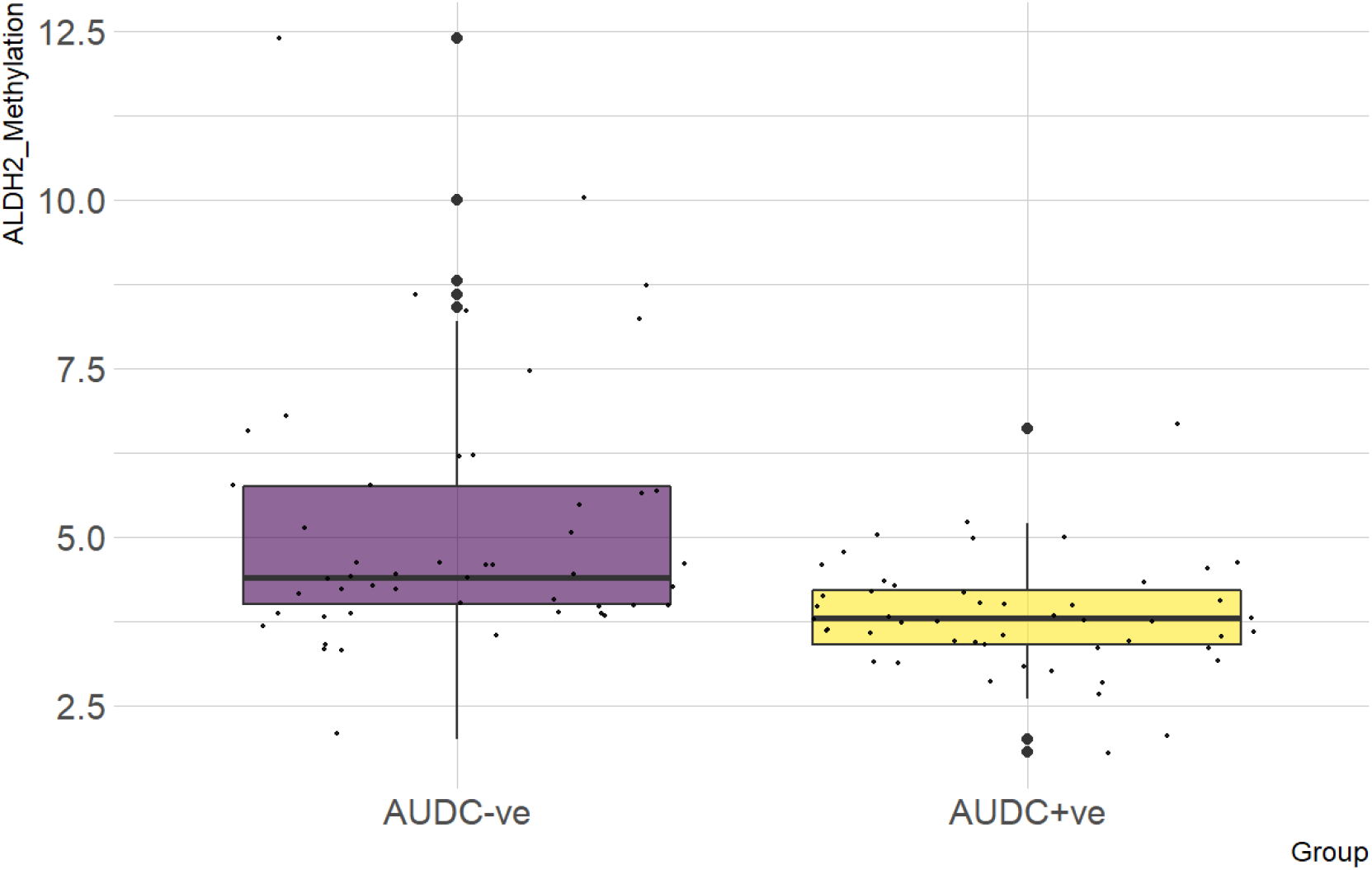
Comparison of mean ALDH2 methylation levels in individuals with AUD with and without cirrhosis. The results indicate that individuals with AUD and cirrhosis had significantly lower mean ALDH2 methylation (p < 0.001) compared to individuals with AUD without cirrhosis. Legend: AUDC-ve – Individuals with AUD without cirrhosis, AUDC+ve – Individuals with AUD with cirrhosis

## Discussion

Our study findings demonstrated that AUD patients who developed cirrhosis, started drinking at a later age, and on an average, were drinking lesser amounts, than those without cirrhosis.

We looked into genotype and allele frequency differences of the *ALDH2* rs2238151 polymorphism between the groups. The frequency of the ‘T’ allele, was higher in the AUDC+ve group than the AUDC-ve group, but this was not statistically significant. Our study investigated differences in global LINE 1 DNA methylation and gene specific *ALDH2* DNA methylation in patients with AUD as a function of liver cirrhosis. LINE 1 DNA methylation did not differ significantly between groups. *ALDH2* CpG methylation was significantly lower in individuals with cirrhosis and lower levels of methylation were seen in this group, in the presence of a T allele. On examining the effect of the rs2238151 allele on methylation levels in the whole set of hundred samples or the AUDC-ve separately, we did not see any effect of the T allele. Thus, suggesting that it is not only a higher T allele frequency in the AUDC+ve group that mediates the lower ALDH2 methylation.

There have been a number of studies which have investigated the role of *ALDH2* gene polymorphisms in individuals with ARLD. Studies from different parts of the world have focused on the role of rs671 in ARLD. A previous study from Eastern India studied three other intronic SNPs including rs2238151 and reported no significant differences in minor allele frequencies between cases with cirrhosis and healthy controls (Roy et al., 2016). Our study also found similar allele frequencies in both our AUD study groups at this SNP and no association with either allele or genotype frequency was seen with cirrhosis in the AUD subjects.

The role of LINE 1 DNA methylation in the etiology of cirrhosis is understudied. An *in vitro* approach using quiescent and early culture-activated hepatic stellate cells (HSCs) showed a global demethylation during activation of HSCs (El Taghdouini et al., 2015). This was replicated in an *in vivo* early-stage animal model of liver fibrosis, where DNA hypomethylation in fibrogenic genes was seen from the onset of fibrosis (Komatsu and Sasaki, 2014). Studies which have compared hepatic tissue from individuals with hepatocellular carcinoma (HCC), cirrhosis and normal liver tissue have shown that only those with HCC exhibited lower global methylation levels (Zheng et al., 2019). We found no differences in LINE 1 DNA methylation measured in peripheral blood of AUD individuals with cirrhosis. This probably points to the fact that a change in global DNA methylation, may represent a critical triggering event only on the onset of HCC.

In the current study we found hypomethylation of ALDH2 gene in AUD with cirrhosis (3.8±0.8) compared to AUD without cirrhosis (5.1±1.9). In a previous study we had reported ALDH2 methylation in a set of controls (4.2±0.7) (Soundararajan et al., 2021). Thus, cirrhosis and HCC (which is often its sequel) both appear to be associated with hypomethylation while the non-cirrhotic phenotype of AUD has a higher methylation level. A higher methylation level might be indicative of lower expression of the ALDH2 and consequent accumulation of acetaldehyde.

We also observed that individuals with a T-allele of rs2238151 had lower methylation levels compared to those with the C-allele in the AUD with cirrhosis group. This kind of interaction between ALDH2 genetic variants and DNA methylation has also been previously described in two studies in individuals with AUD. These studies investigated the influence of the ALDH2 promoter polymorphism rs886205 on DNA methylation and found that DNA methylation levels differed between those with A and G alleles and that methylation changes during the course of alcohol withdrawal were also distinct (Haschemi Nassab et al., 2016; Pathak et al., 2017). While our sample size for this type of analysis was small, it highlights the importance of looking into genetic influences on DNA methylation.

ALDH2 metabolizes a number of substrates in addition to acetaldehyde including lipid peroxidation derived compounds such as 4-hydroxy-2-nonenal, malondialdehyde and 3,4-dihydroxyphenylacetaldehyde. Hepatotoxicity as seen in ARLD and neurodegeneration as seen in Alzheimer’s Disease and Parkinson’s disease may be a result of competition between acetaldehyde and these compounds for ALDH2 mediated metabolism (Marchitti et al., 2008). As a result, *in vivo* animal studies using hepatic ALDH2 knockout mice have shown a reduced preference for heavy alcohol consumption, which may limit systemic side effects (Guillot et al., 2019). Further, another study found that a selective small-molecule ALDH2 activator, N-(1,3-benzodioxol-5-ylmethyl)-2,6-dichlorobenzamide (Alda-1), accelerates aldehyde clearance and reverses hepatic steatosis and apoptosis in mice through pharmacological activation of ALDH2 (Zhong et al., 2015).

Our study did not include functional assays of ALDH2 mRNA or protein levels that could have helped to ascertain the impact of genetic and epigenetic differences on enzyme activity;this may need to be explored in further studies. While the sample size for methylation analysis is modest the current study provides some insight into the prevailing differences in individuals exposed to environmental insults like heavy alcohol use. Since tissues from the liver and brain would be accessible together only post-mortem, it would have limited utility in comparing ongoing tissue specific changes as a consequence of excessive alcohol use. Evaluating the blood epigenome is the ideal option as there is significant evidence to show that brain, blood and liver, share similar methylation trends at most loci. Further studies maybe necessary to elucidate tissue specific mechanisms of alcohol related complications.

## Conclusion

Our study found that DNA methylation at *ALDH2* was significantly lower in AUD individuals with cirrhosis and this was further exacerbated by the occurrence of the T allele in this group of patients. Future studies need to be conducted to determine the functional impact of these changes and also to see if similar changes are also observed in different tissues using cellular models. This will offer more insight into the pathophysiology of alcohol related end organ complications and vulnerability to such complications.

## Data Availability

Data referred to in the manuscript is available upon request from the first/corresponding author

