## Supplementary_Tables for "Genetics and Epigenetics of Aldehyde Dehydrogenase (ALDH2) in Alcohol Related Liver Disease"

Supplementary Table 1: Differences between Genotype and Allele frequency of ALDH2 rs2238151 in individuals with AUD with and without cirrhosis in the DNA methylation subset

| **Gene and SNP** | **Genotype Frequency** | **AUDC-ve**  **N=50**  **n, %** | **AUDC+ve**  **N=50**  **n, %** | **P** | **Chi-Square** |
| --- | --- | --- | --- | --- | --- |
| **ALDH2**  **rs2238151** | CC | 32 (0.64) | 26 (0.52) | 0.4 | 1.4 |
|  | CT | 16 (0.32) | 21 (0.42) |  |  |
|  | TT | 2 (0.04) | 3 (0.06) |  |  |
|  | **Allele frequency** | **2n=100** | **2n=100** | **P** | **Chi-Square** |
|  | C | 0.8 | 0.73 | 0.2 | 1.3 |
|  | T | 0.2 | 0.27 |  |  |

Supplementary Table 2: Differences in average DNA methylation of LINE1 CpG sites in individuals with AUD with and without cirrhosis

| **LINE1 CpG sites** | **Methylation Levels (Mean±SD)** | | **p-value** |
| --- | --- | --- | --- |
|  | **AUDC-ve**  **N=50** | **AUDC+ve**  **N=50** |  |
| CpG1 | 82.7±3.4 | 82.4±4.2 | 0.68 |
| CpG2 | 70.3±6.3 | 69.6±6.1 | 0.55 |
| CpG3 | 72.0±10.3 | 72.5±2.6 | 0.73 |
| CpG4 | 80.2±3.3 | 80.3±3.2 | 0.92 |
| CpG5 | 61.1±3.4 | 60.9±2.4 | 0.76 |
| Average | 73.3±3.9 | 73.0±2.3 | 0.66 |
